# Biomarkers of RV dysfunction in HFrEF identified by direct tissue proteomics: extracellular proteins fibromodulin and fibulin-5

**DOI:** 10.1101/2024.05.16.24307514

**Authors:** Matěj Běhounek, Denisa Lipcseyová, Ondřej Vít, Petr Žáček, Pavel Talacko, Zuzana Husková, Soňa Kikerlová, Tereza Tykvartová, Peter Wohlfahrt, Vojtěch Melenovský, Jan Beneš, Jiří Petrák

## Abstract

**BACKGROUND:** Right ventricular dysfunction (RVD) is a commonly accompanying condition in patients with heart failure with reduced ejection fraction (HFrEF), and is associated with poor prognosis. However, no specific biomarker of RVD is available for clinical routine.

**METHODS:** We performed a label-free proteomic analysis of myocardium from the left and right ventricles of HFrEF patients with and without RVD who underwent heart transplantation (n=20). Concentrations of two extracellular matrix proteins with highest myocardial upregulation in RVD – fibromodulin (FMOD) and fibulin-5 (FBLN5) - were assayed in the blood, compared to controls (n=65) and tested in separate large cohort of HFrEF patients (n=232) to test association of the two proteins with RV function and long-term outcome.

**RESULTS:** FMOD and FBLN5 plasma concentrations progressively increased with worsening RV function (p = 0.0001 and 0.004), while there was no association with LV dysfunction, BMI or kidney function. In survival analysis, plasma levels of both FMOD and FBLN5 were significantly associated with patient outcome (p = 0.005 and 0.0004). Using reclassification analysis, addition of plasma FBLN5 to right ventricle function assessment led to an improvement in AUC after 4 years compared to RV function assessment only. Similarly, adding plasma fibromodulin to the multi-parametric MAGGIC score showed a significantly higher AUC after 4 years. Importantly, the prognostic value of both biomarkers was additive.

**CONCLUSION:** Our study proposes that circulating levels of FMOD and FBLN5 may serve as a new biomarkers of RVD in HFrEF patients.

## Introduction

Right ventricular dysfunction (RVD) is a major complicating condition of heart failure (HF) with both preserved and reduced EF and is associated with poor prognosis (1–3). Although no specific RV dysfunction-targeted therapy is currently available, RV dysfunction is commonly present in advanced HF, and may herald clinical worsening with the need of intensification or therapy or timely indication to heart transplantation. Development of severe RV dysfunction may preclude use of long-term left ventricular assist devices (LVAD), limiting options for long-term survival (4).

RV function is currently assessed mainly with imaging, and echocardiography is the most commonly employed method. There is no specific circulating biomarker available in clinical practice to recognize specifically RVD in the context of HFrEF. Identification of a biomarker specific for RVD could add to echocardiographic RV evaluation, better risk-stratify patients and help further decision-making in advanced HF management.

Right ventricle (RV) has a different embryological origin than left ventricle (LV) and operates in different pressure conditions. Dysfunction of the right heart may result from- or translate into specific proteome changes in both ventricles. The proteins with altered expression can be assistive as disease markers, pending their altered expression is mirrored but their altered concentration in blood. Such proteins can be either passively released from myocardium as a result of tissue leakage caused by cell death or injury, or actively secreted from the cells to the interstitial fluid. To look for such potential markers of RVD in patients with advanced HF we performed a label-free proteomic analyses of myocardium from both ventricles of HFrEF patients with and without RVD. Among the differentially expressed proteins we identified significant upregulation of two secreted proteins of extracellular matrix (ECM) - fibromodulin (FMOD, upregulated in RV) and fibulin 5 (FBLN5, upregulated in LV) in the heart of HFrEF patients with RVD. Their upregulation was mirrored in increased concentrations in blood plasma. We have evaluated the clinical utility of the two promising candidate biomarkers in a large cohort of advanced HFrEF patients.

## METHODS

All methods are described in detail in the **Supplementary Material.**

### Patients

Subjects with symptomatic but hemodynamically stable HFrEF (LVEF <40%) that had a duration of at least 6 months were enrolled in the study (1, 5). Those with potentially reversible left-ventricular (LV) dysfunction (planned valve surgery, revascularization, or tachycardia-induced cardiomyopathy) were excluded. Echocardiography was performed in all patients upon enrollment. All research was performed in accordance with relevant guidelines/regulations, the protocol was approved by the Ethics Committee of the Institute for Clinical and Experimental Medicine, and all subjects signed an informed consent.

### Patients cohort # 1 - Proteomics cohort

A cohort of 122 consecutive patients that underwent heart transplantation without preceding use of LVAD (to exclude the effect of unloading) was divided according to the RV function measured by fractional area change (FAC) measured by a single experienced echocardiographer on echocardiography scan prior heart transplantation. A subgroup of 10 HFrEF male patients with severe RV dysfunction (1st quartile of FAC), and 10 age, body size, etiology and comorbidity-matched HFrEF male patients with preserved RV function (4th quartile of FAC) were used for proteomic analysis. In addition, 10 age and body size matched subjects without a clinical history of HF and with normal both LV and RV function that were refused as heart donors due to hemodynamic instability or moderate coronary lesions were added to the analysis as a proxy for non-failing controls. Tissue samples of right and left heart ventricle free wall were snap-frozen and stored at -80°C immediately after the heart explantation. Samples were later pulverized in liquid nitrogen using a mortar and pestle, aliquoted and stored at -80°C for proteomic analysis and immunoblotting. Patient data for the proteomics cohort are provided in **Table 1**.

**Table 1.**
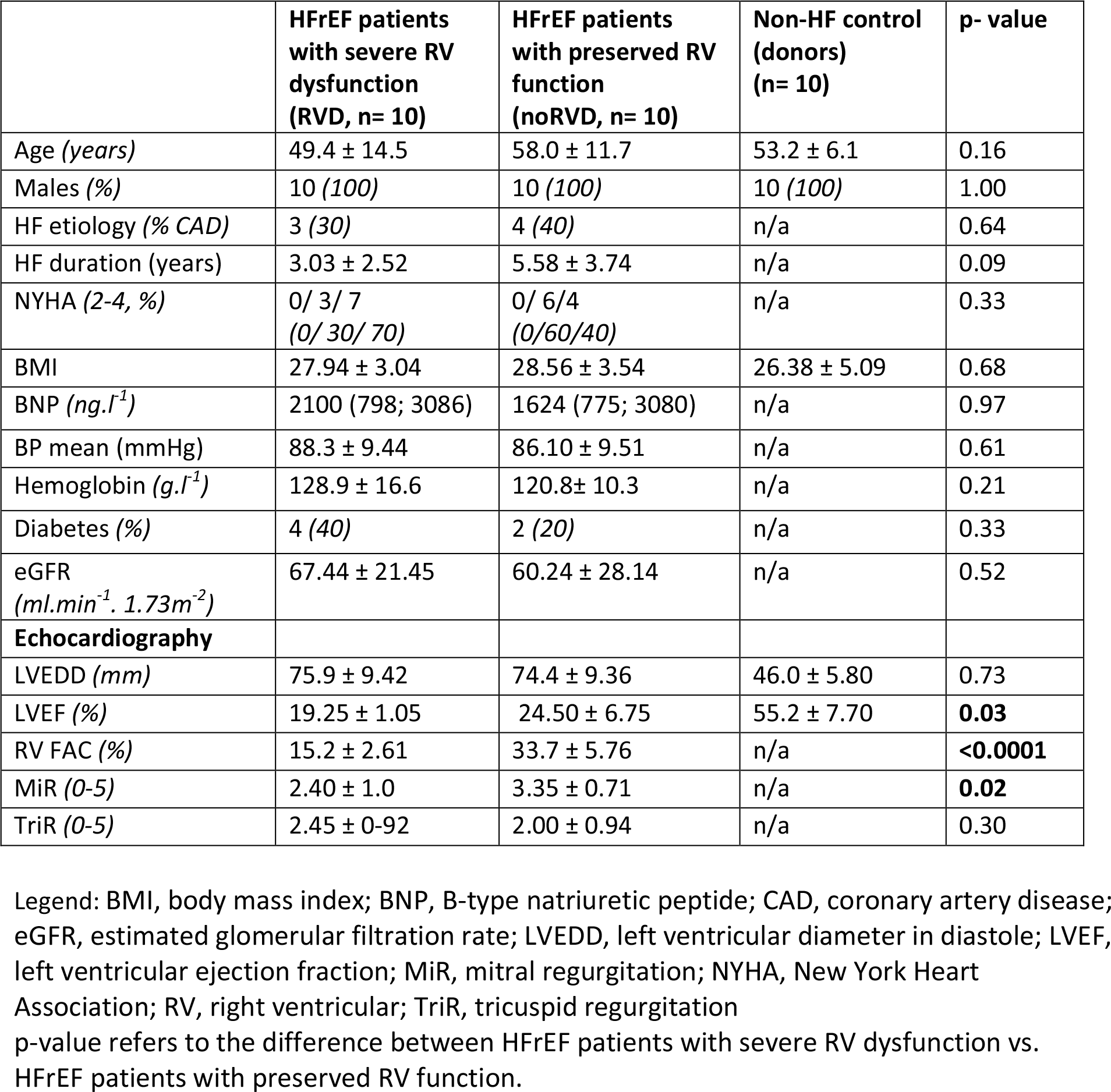
HFrEF Patients with and without RVD and controls selected for proteomic analysis.

**Table 2A.** Proteins differentially expressed in the right ventricle of HFrEF patients with RVD compared to HFrEF patients without RVD.

| Accession | Gene name | Protein name | RIGHT VENTRICLE |  |  |
| --- | --- | --- | --- | --- | --- |
|  |  |  | Unique peptides | RVD/noRVD fold-change | p-value |
| Upregulated in RVD |  |  |  |  |  |
| P35580 | MYH10 | Myosin-10 | 38 | 3.61 | 0.0113 |
| Q06828 | FMOD | Fibromodulin | 6 | 3.35 | 0.0141 |
| P55083 | MFAP4 | Microfibril-associated glycoprotein 4 | 4 | 2.76 | 0.0017 |
| P09936 | UCHL1 | Ubiquitin carboxyl-terminal hydrolase isozyme L1 | 15 | 2.76 | 0.0022 |
| Q9ULD0 | OGDHL | 2-oxoglutarate dehydrogenase-like, mitochondrial | 13 | 2.70 | 0.0154 |
| Q9NZM1 | MYOF | Myoferlin | 36 | 2.46 | 0.0021 |
| P35443 | THBS4 | Thrombospondin-4 | 15 | 2.43 | 0.0005 |
| P13611 | VCAN | Versican core protein | 8 | 2.38 | 0.0041 |
| P21810 | BGN | Biglycan | 8 | 2.19 | 0.0380 |
| P50225 | SULT1A1 | Sulfotransferase 1A1 | 6 | 2.08 | 0.0090 |
| Downregulated in RVD |  |  |  |  |  |
| O14896 | IRF6 | Interferon regulatory factor 6 | 6 | 0.35 | 0.0346 |
| Q96C11 | FGGY | FGGY carbohydrate kinase domain-containing protein | 3 | 0.48 | 0.0183 |

**Table 2B.** Proteins differentially expressed in the left ventricle of HFrEF patients with RVD compared to HFrEF patients without RVD.

| Accession | Gene name | Protein name | LEFT VENTRICLE |  |  |
| --- | --- | --- | --- | --- | --- |
|  |  |  | Unique peptides | RVD/noRVD fold-change | p-value |
| Upregulated in RVD |  |  |  |  |  |
| Q9UBX5 | FBLN5 | Fibulin-5 | 6 | 2.47 | 0.0032 |
| Q9UBG0 | MRC2 | C-type mannose receptor 2 | 11 | 2.38 | 0.0162 |
| O94788 | ALDH1A2 | Retinal dehydrogenase 2 | 13 | 2.15 | 0.0102 |
| Downregulated in RVD |  |  |  |  |  |
| Q96QA5 | GSDMA | Gasdermin-A | 9 | 0.33 | 0.0305 |
| Q96LJ7 | DHRS1 | Dehydrogenase/reductase SDR family member 1 | 4 | 0.38 | 0.0041 |
| Q5XPI4 | RN123 | E3 ubiquitin-protein ligase RNF123 | 12 | 0.47 | 0.0385 |

### Patients cohort # 2 - Clinical cohort

A group of 232 HFrEF patients referred to IKEM for evaluation of advanced therapies or implantable cardioverter-defibrillator (ICD) implantation and 65 non-HF controls was used for this analysis. Patients were prospectively enrolled between 2008 and 2016 and underwent complex in-hospital evaluation, including echocardiography and blood sampling. Patients were prospectively followed for the occurrence of an adverse outcome that was defined as the combined endpoint of death, urgent heart transplantation, or ventricular assist device implantation. Because the time to non-urgent transplantation reflects donor availability rather than recipient’s condition, patients who received a non-urgent heart transplant were censored as having no outcome event at the day of transplantation. The control group consisted of 65 subjects (36 subjects without the evidence of coronary artery disease or heart failure evaluated for the presence of patent foramen ovale), 29 subjects were healthy hospital employees. Patient data for the clinical cohort are provided in **Table 3**.

**Table 3.**
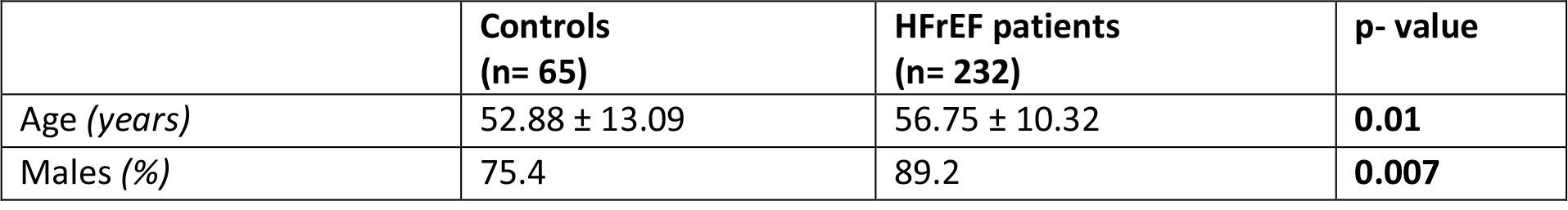
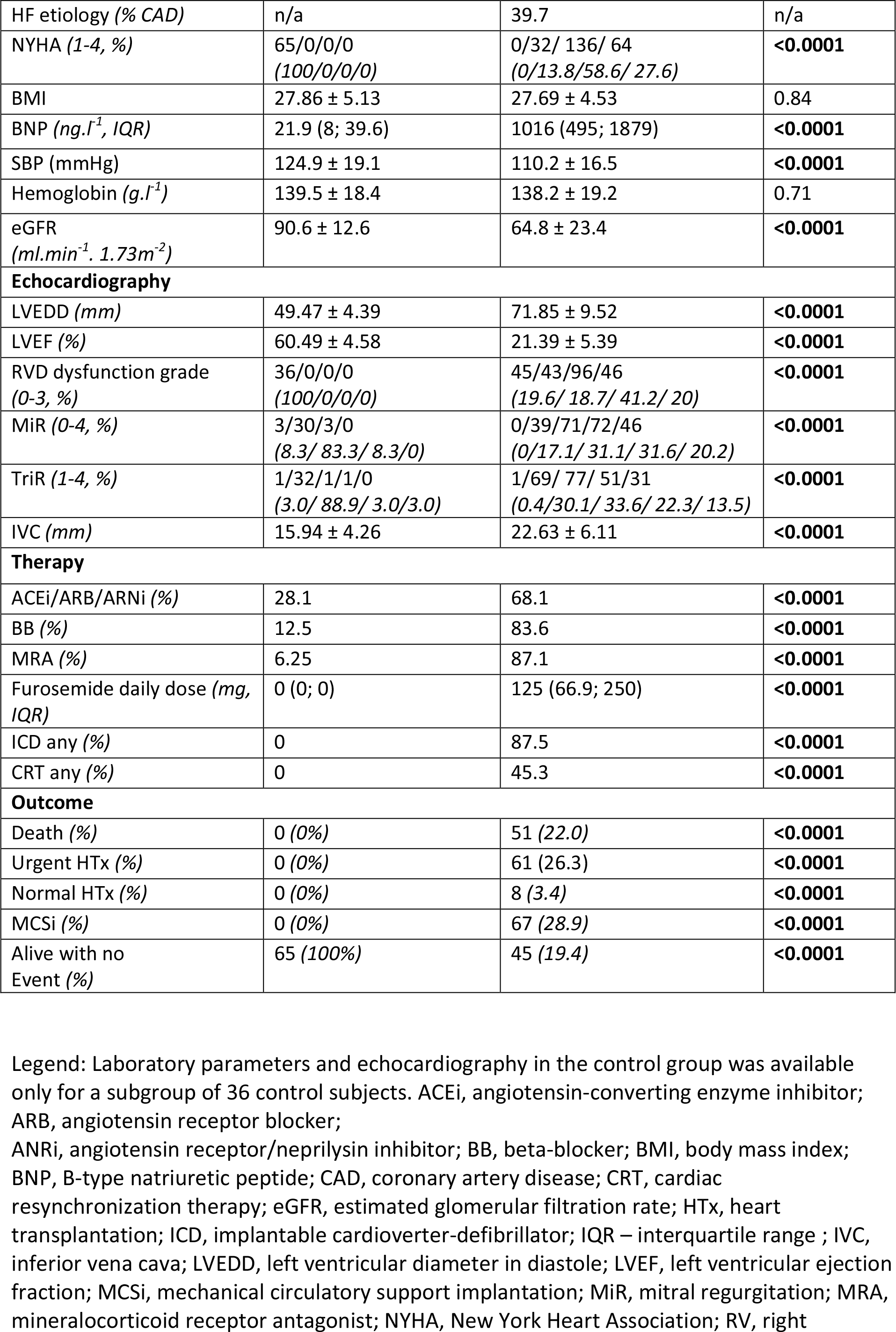
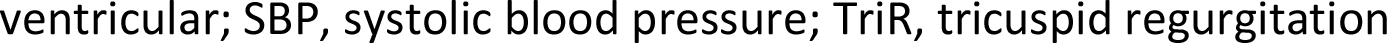
Patients characteristics (Clinical cohort).

## RESULTS

### Proteomic analysis

Myocardial samples of left and right ventricles from 10 patients with HFrEF and severe RV dysfunction (RVD), 10 patients with HFrEF and preserved RV function (noRVD) and 10 control individuals were subjected to quantitative proteomic analysis. Since RVD may be associated also with molecular changes in the left ventricle, and in respect for their different ontology, wall thickness, working pressures, physiology and pathology, we analyzed separately both right (RV) and left ventricles (LV). The description of patients and controls is given in **Table 1**.

The label-free quantification (LFQ) proteomic analyses of RV and LV resulted in identification of 3477 and 3633 protein groups, respectively. Supervised discrimination analysis sPLSDA (sparse partial least squares discrimination analysis) of the proteomic data from RV a LV clearly separated non-failing (control) hearts from the failing hearts and, albeit less completely, also samples from RVD patients with (red) and noRVD patients (green) in both ventricles (**Figure 1**).

**Figure 1.**
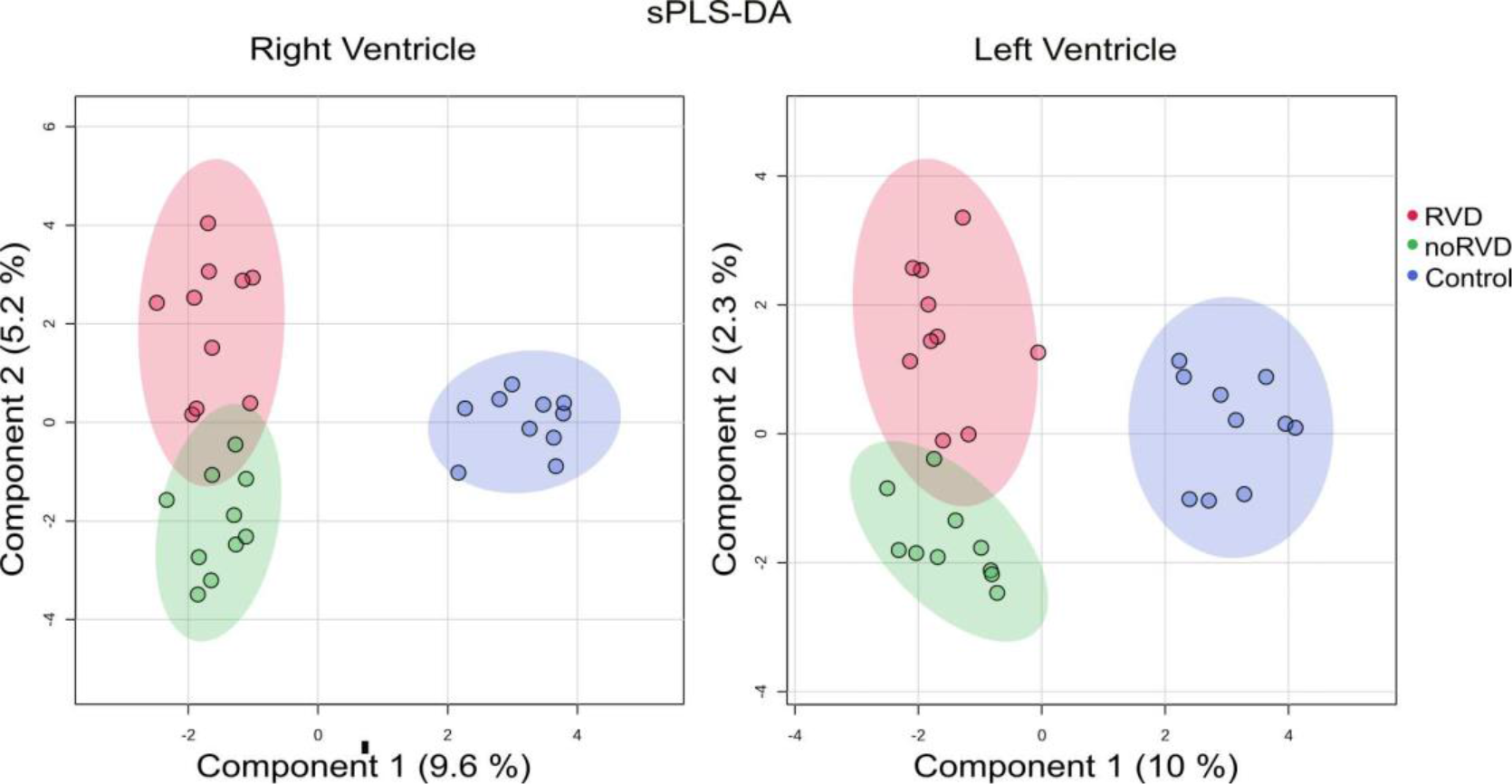
Supervised discrimination analysis sPLS-DA (sparse least squares discrimination analysis) of the quantitative proteomic data. Results of the separate analyses of right and left ventricle is shown.

This incomplete separation of RVD and noRVD proteomic profiles reflects rather mild changes in myocardial proteome of HFrEF patients with (RVD) and without RVD (noRVD). In fact, we identified only 12 and 6 differentially expressed proteins (>2-fold, p-val 0.05) in the right and left ventricles of RVD patients, respectively, compared to noRVD patients (**Figure 2**). None of the 19 RVD- associated changes in protein expression was identified simultaneously in both ventricles. **Tables 2A and 2B** list the identified differentially expressed proteins. List of all identified proteins is available as a **Supplementary table 1.** The mass spectrometry data have been deposited to the ProteomeXchange Consortium via the PRIDE partner repository with the dataset identifier PXD043768.

**Figure 2.**
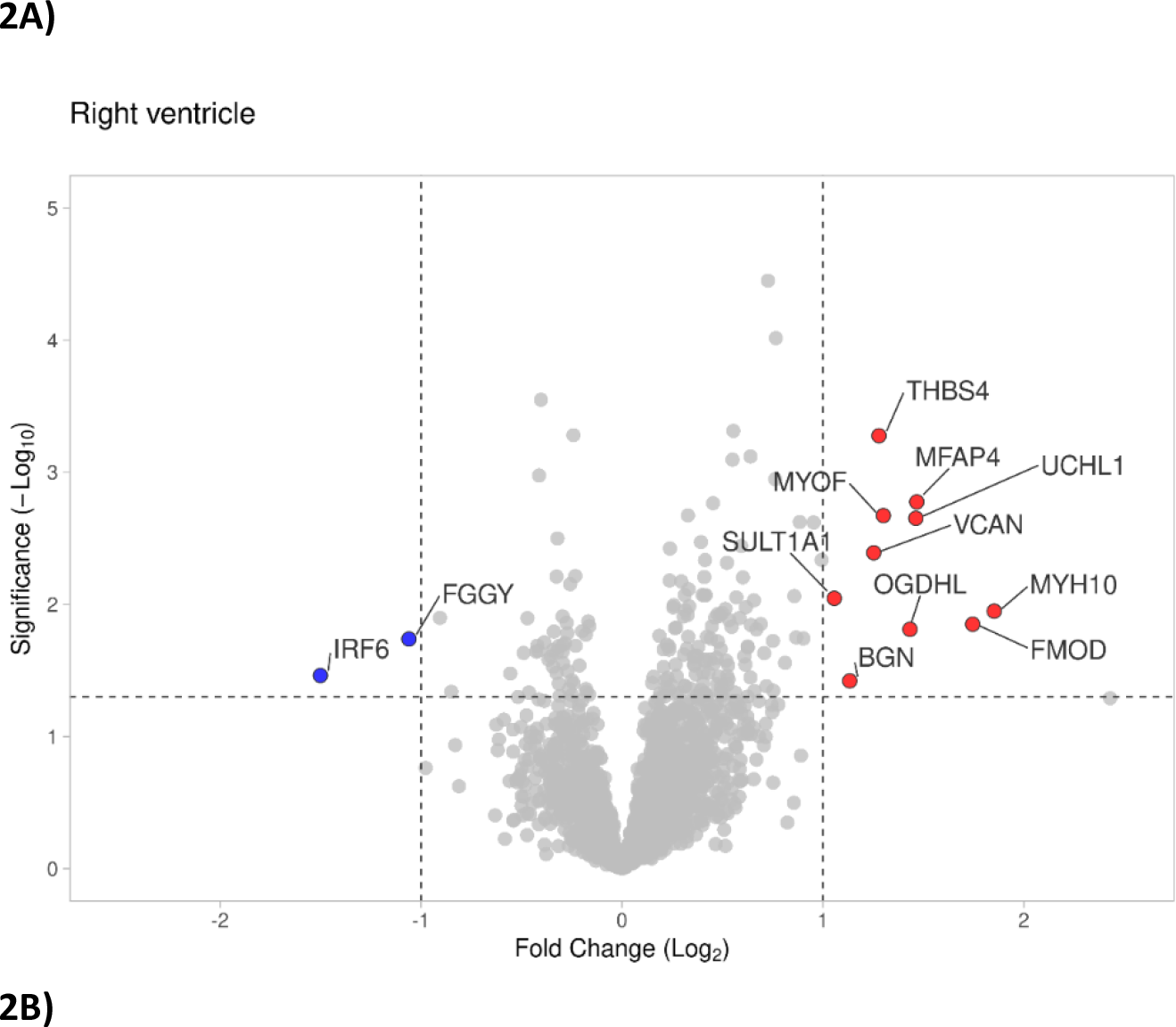
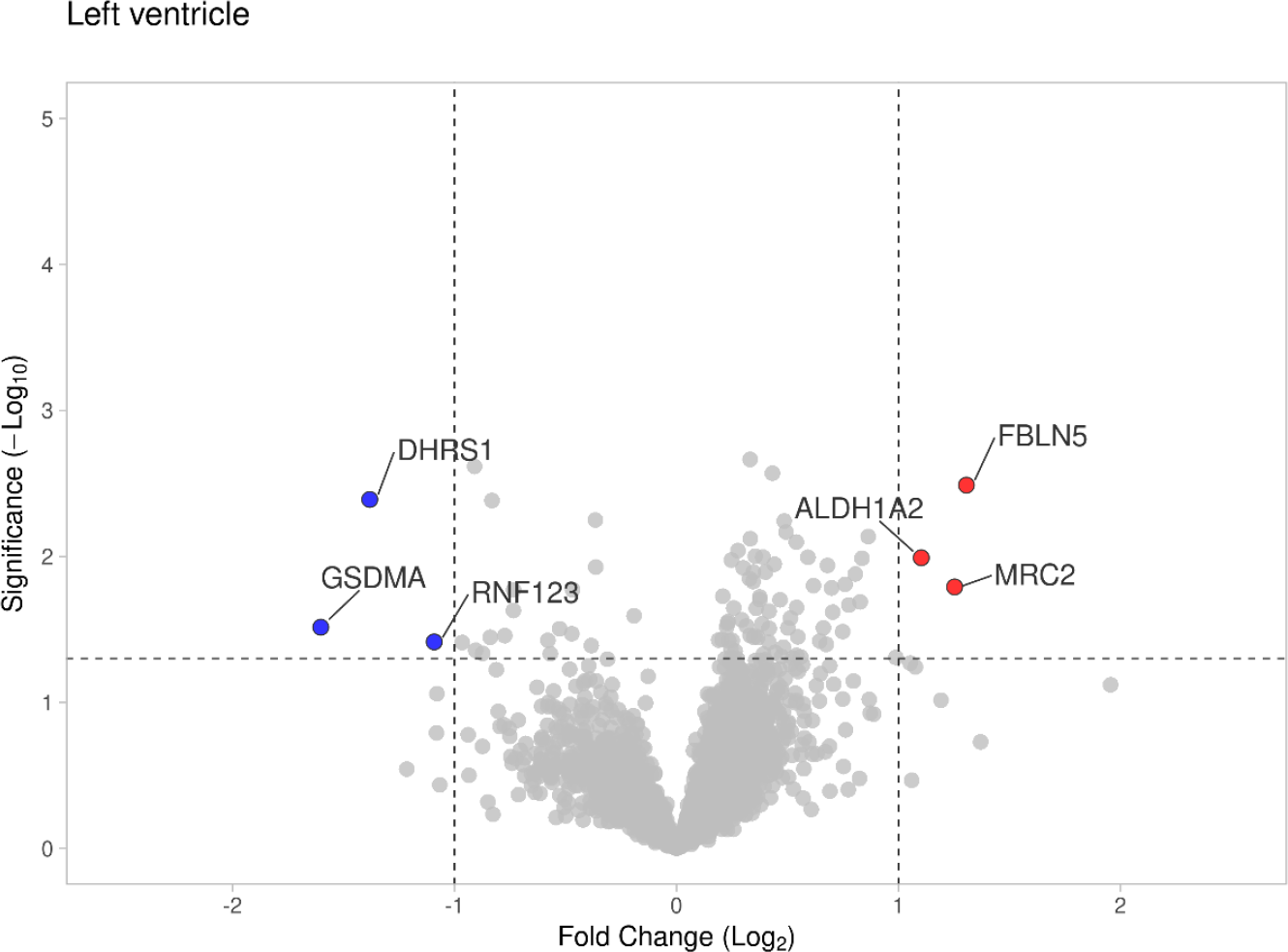
LFQ proteomic analyses of ventricular myocardium of HFrEF patients with RVD versus noRVD. Proteins upregulated in RVD versus noRVD are shown in red, downregulated in blue. Threshold fold- change >2-fold, Adjusted p-value <0.05). Gene names are shown instead of the full-length protein names. **A)** Protein expression changes in the right ventricle. **B)** Protein expression changes in the left ventricle

### Bioinformatic analysis – Pathway enrichment

Since the proteome changes were limited and the numbers of the differentially expressed (>2-fold) proteins were low, gene ontology (GO) pathway analysis revealed significant enrichment only among the proteins upregulated in right ventricle (**Supplementary figure 1**). “ECM matrix organization” (adj. p-val <0.001) was the process with the highest number of matched proteins. No enrichment was indicated in LV, neither among the downregulated RV proteins.

In agreement with the GO enrichment analysis, 5 out of the 10 proteins markedly upregulated in the RV of the of the RVD group are ECM proteins annotated as “secreted, extracellular space, extracellular matrix” according to the UniProt database, namely fibromodulin (FMOD), microfibril-associated glycoprotein 4 (MFAP4), versican (VCAN), biglycan (BGLN) and thrombospondin 4 (THBS4). In the left ventricle of the RVD group, the most markedly upregulated protein was fibulin-5 (FBLN5) which is also annotated as secreted/ECM protein. Additionally, three other proteins upregulated in it the hearts of patients with RVD, namely (non-muscle) myosin 10 (MYH10), ubiquitin carboxy-terminal hydrolase isoenzyme L1 (UCHL1) and C-type mannose receptor are known to participate in the process of ECM remodeling as well, despite their intracellular localization (6–8). The results of our proteomic analysis thus clearly pointed toward pronounced ECM remodeling as a hallmark of RV dysfunction in HFrEF patients.

### Confirmation of the proteomic data in myocardial tissue

In order to provide an independent confirmation of the proteomic data, we verified relative protein expression of six proteins identified as differentially expressed in heart of HFrEF patients with RVD, using specific antibodies. The same samples were used for both the proteomic and the confirmatory analysis. The results were in full agreement with the proteomic analysis expression of FMOD, MFAP4, UCHL1, MYOF and VCAN was confirmed as significantly differentially upregulated in the right ventricle, while FBLN5 was in significantly upregulated in the LV of HFrEF patients with RVD compared to those with preserved RV function. All the proteins were also significantly upregulated relative to respective ventricles of non-failing controls. (**Figure 3**).

**Figure 3.**
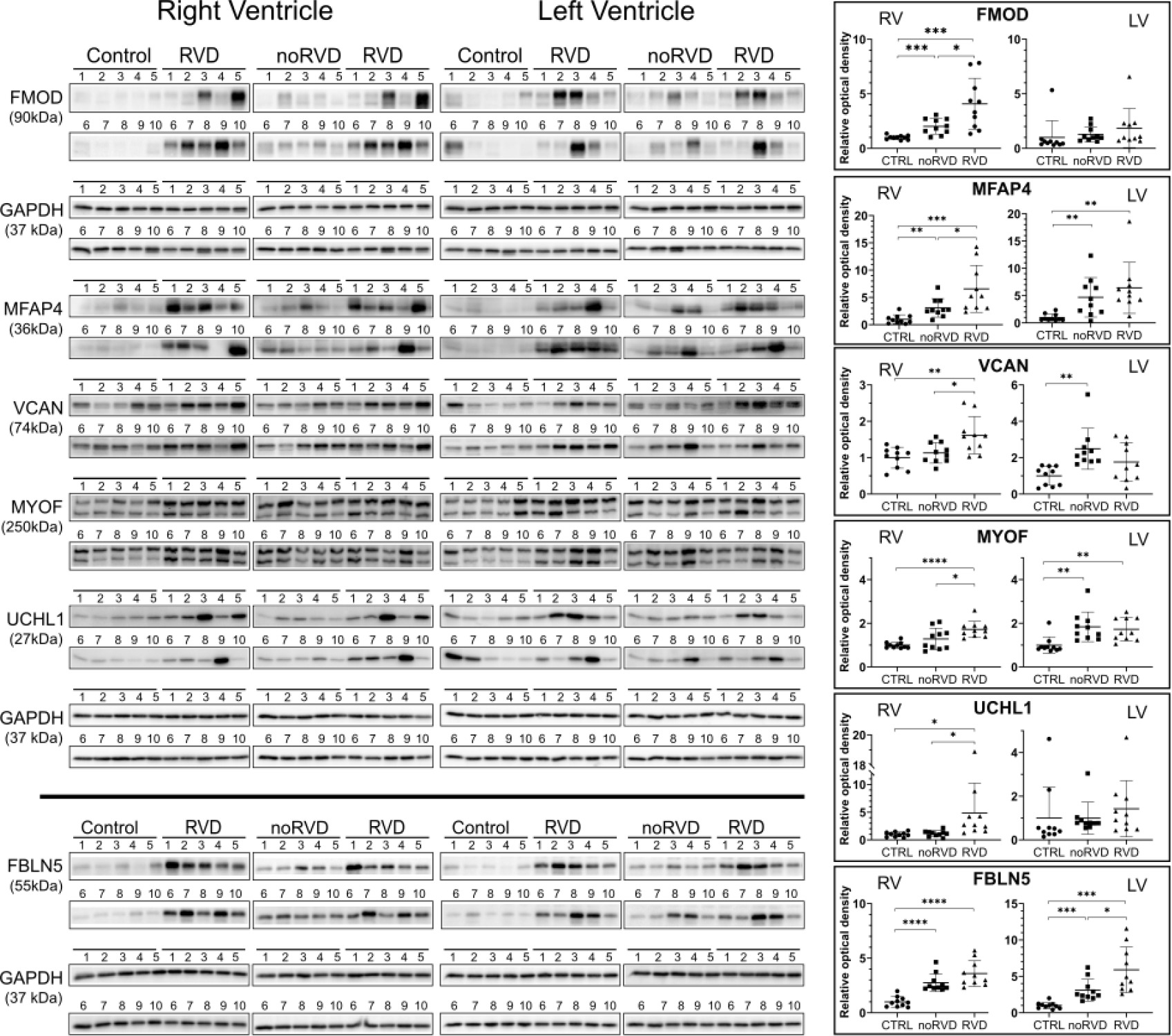
**Confirmation of differential expression by western blotting. A**) Relative protein expression was determined in both ventricles of all the patients included in the proteomic analysis (RVD and noRVD). To obtain also the information on the “background” expression, non-failing donor heart samples were included (Control, CTRL). RV and LV were analyzed separately; GAPDH was used as a loading control. **B**) Densitometry of the western blots. Relative optical density of the bands normalized to the controls is plotted. Statistical significance is indicated where reached. Gene names are shown instead of full protein names FMOD (fibromodulin), MFAP4 (microfibril-associated glycoprotein 4), VCAN (versican), MYOF (myoferlin), UCHL1 (ubiquitin carboxy-terminal hydrolase isoenzyme L1), FBLN5 (fibulin5).

### Blood detection of secreted myocardial proteins linked to the presence of RV dysfunction

Five out of 10 proteins upregulated in the RV of patients with RVD are secreted proteins - components of extracellular matrix (ECM), and we reasoned that secreted proteins could serve as circulating biomarkers. Among them fibromodulin (FMOD) is the most markedly (3.3-fold according the proteomic analysis) upregulated ECM protein in the right ventricle of HFrEF patients with RVD. In the left ventricle of HFrEF patients with RVD, fibulin-5 (FBLN5) is the most strongly upregulated protein (2.5-fold) and the only upregulated secreted ECM molecule. Looking for RVD markers, we therefore asked whether the marked myocardial upregulation of these myocardial secreted/ECM proteins is mirrored in the blood plasma of HFrEF patients with RVD. Proteins from both ventricles were considered since both ventricles may be a source of (potentially distinct) information on RVD.

### Fibromodulin and fibulin-5 in the blood plasma

Using specific sandwich ELISA, we first determined plasma concentration of FMOD and FBLN5 in the blood samples of the 20 HFrEF individuals (collected at the time the transplantation) included in the proteomic analysis (the cohort #1) and healthy male volunteers of comparable age (n=10). Mean plasma FMOD concentrations were 17.58 (SD 3.84), 22.19 (SD 8.18) and 27.14 (SD 8.86) ng/mL in controls, noRVD and RVD patients with HFrEF, respectively. Mean plasma concentrations of FBLN5 reached 54.32 (SD 15.78), 71.63 (SD 23.61) and 96.32 (SD 48.08) ng/mL in the same groups (**Supplementary figure 2**). In agreement with the increased myocardial expression, plasma concentration of both proteins in RVD group were higher compared to controls (p= 0.005, and p=0.017) and tended to be numerically higher compared to noRVD group (Supplementary figure 2). Considering the limited size of the original cohort #1 we evaluated the plasma concentration of FMOD and FBLN5 in a large cohort of well-characterized patients with advanced HF.

### The utility of fibulin-5 and fibromodulin in clinical scenario

We tested the utility of fibulin-5 and fibromodulin plasma concentrations in a cohort of 232 HFrEF patients and 65 controls. Patients (56.75 ± 10.32 years, 89.2% males) presented with advanced HF (86.2% NYHA III/IV, LVEF of 23.3%) and significant number of patients (n= 142, 61.7%) had moderate or severe RV dysfunction (details in **Table 3).** Fibromodulin and fibulin-5 plasma levels progressively and significantly increased with worsening RV function (**Figure. 4**).

**Figure 4.**
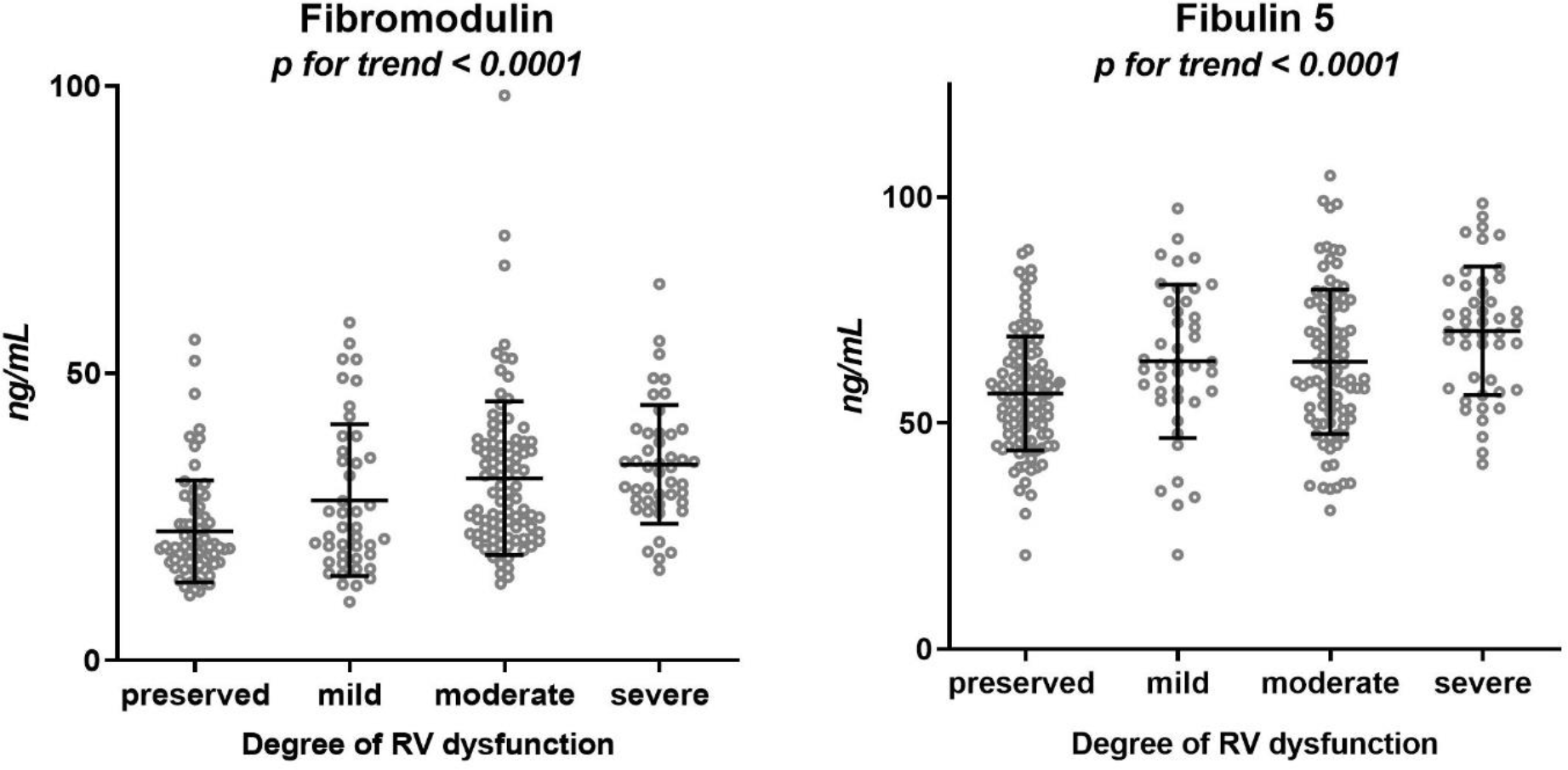
The plasma concentrations of FMOD and FBLN5 with respect to RV dysfunction. Patients with increasing level of RV dysfunction presented with increasing levels of both FBLN5 and FMOD. Mean and standard deviation are shown for each group. The assessment of RV function was performed as described in methods. Preserved indicates preserved (normal) RV function; mild, moderate, severe indicate mild, moderate and severe RV dysfunction, respectively.

### Regression analysis

We have further analyzed what variables were associated with fibromodulin and fibulin-5 plasma levels. In HFrEF patients, a stepwise linear regression showed that fibromodulin level was significantly associated with RVEF only (p< 0.0001, RVEF explains 7.4% of fibromodulin variability). Similarly, a significant association between fibulin-5 and RVEF (p=0.004, RVEF explains 3.2% of fibulin-5 variability) and age (p<0.0001, age explains 7.1% of fibulin-5 variability) was found. There was no association of fibulin-5 or fibromodulin plasma levels with LVEF, BMI or kidney function (eGFR).

### Fibulin-5 and fibromodulin as predictors of adverse outcome in HFrEF

During a follow up of a median period of 4.4 years (IQR 3.6; 5.0), 179 patients (77.2%) in our clinical cohort had an adverse outcome defined as death/urgent heart transplantation/implantation of mechanical circulatory support. Cox proportional hazard model revealed that both FMOD and FBLN5 were significantly associated with patient outcome, with hazard ratio (HR) of 1.20 (95% CI 1.05- 1.37), p= 0.005 per one standard deviation (SD) of FMOD plasma concentration and HR of 1.30 (95% CI 1.12- 1.51), p= 0.0004 per one SD of FBLN5 concentration.

### Clinical value of fibromodulin and fibulin-5 plasma levels in addition to RV function assessment

As both candidate markers FMOD and FBLN5 associated with patient outcome and showed a significant association with RV function, we have asked whether any of the proteins could improve the prognostic stratification beyond RV function assessment by echocardiography. When adjusted for RV dysfunction grade, FMOD (upregulated in RV) was no longer a significant predictor of outcome in Cox regression analysis (HR 1.29, 95% CI 0.98- 1.30, for each SD of FMOD concentration, p= 0.08).

Similarly, the analysis of area under the curve (AUC) showed no improvement found for a combination of RV-dysfunction grade with FMOD at a single time point of 4 years, (176 out of a total of 179 outcome events were reached at this time point). In contrast, FBLN5 (upregulated in LV) remained significantly associated with outcome even when adjusted for RV dysfunction grade (HR 1.30, 95% CI 1.12- 1.51 for each SD of FBLN5 concentration, p= 0.0006). In addition, there was a significant improvement in AUC for RV-dysfunction grade combined with fibulin-5 level compared to RV dysfunction grade at 4 years of follow-up **(Table 4).**

**Table 4.** The comparison of RV function assessment and MAGGIC score with addition of FBLN5 and FMOD. Assessment of RV function. Addition of FMOD to the RV function assessment by echocardiography did not significantly improved AUC. However, addition of FBLN5 lead to an improvement in AUC compared to RV function assessment only. Net reclassification improvement (NRI) was significant for the overall score (NRI) and for reclassification of patients with events into higher-risk group (NRI+) but not for the reclassification of patients without an event into a lower-risk group (NRI-). MAGGIC score. Adding FMOD to MAGGIC score showed a significantly higher AUC, while addition of FBLN5 to the MAGGIC score did not change the AUC significantly. However, both FBLN5 and FMOD significantly improved the overall NRI score as well as the reclassification of patients with events into higher-risk group (NRI+). The reclassification of patients without an event into lower-risk group (NRI-) was significant for FMOD only.

| Area under the curve (AUC) |  |  |  |  | NRI |  |  |
| --- | --- | --- | --- | --- | --- | --- | --- |
| RV dysf. grade | RV dysf. grade + FBLN5 | Delta AUC | 95% CI | p- value | NRI (95%CI) | NRI+ (95%CI) | NRI- (95%CI) |
| 0.570 | 0.653 | 0.083 | 0.01; 0.166 | **0.049** | 0.41 (0.13; 0.63) | 0.17 (0.08; 0.31) | 0.24 (-0.02; 0.35) |
| MAGGIC | MAGGIC + FMOD | Delta AUC | 95% CI | p- value | NRI (95%CI) | NRI+ (95%CI) | NRI- (95%CI) |
| 0.579 | 0.658 | 0.079 | 0.024; 0.134 | **0.005** | 0.37 (0.06; 0.80) | 0.12 (0.01; 0.21) | 0.25 (0.02; 0.61) |
| MAGGIC | MAGGIC + FBLN5 | Delta AUC | 95% CI | p- value | NRI (95%CI) | NRI+ (95%CI) | NRI- (95%CI) |
| 0.579 | 0.650 | 0.071 | -0.005; 0.147 | 0.07 | 0.33 (0.12; 0.65) | 0.15 (0.08; 0.26) | 0.18 (-0.04; 0.45) |

We have further employed net reclassification improvement (NRI) index that attempts to quantify how well a new model reclassifies subjects - either appropriately or inappropriately - as compared to an old mode (9). FBLN5 markedly improved the continuous NRI, with a significant improvement in event NRI (NRI+), but without change in nonevent NRI (NRI-).

### Adding FMOD and FBLN5 to MAGGIC score improves long-term prediction

In clinical practice, the prognosis of HF patients is often evaluated by composite scoring systems that help to guide therapeutic decisions. MAGGIC score is one of those and encompasses various patient characteristics, degree of LV dysfunction (LV-ejection fraction), HF pharmacotherapy and comorbidities (10). We evaluated whether plasma concentrations of FMOD or FBLN5 add any additional prognostic information beyond MAGGIC score (**Table 4**). Adding plasma FMOD to MAGGIC score showed a significantly higher AUC after 4 years (p= 0.005). In addition to that, FMOD correctly reclassified both patients with event into more high-risk group (NRI+) as well as those without event into lower-risk group (NRI-). Adding plasma FBLN5 to the MAGGIC score numerically increased AUC, however the change was not statistically significant (p= 0.07). However, FBLN5 correctly reclassified patients with events into more high-risk group (significant NRI+).

### A combined prognostic role of FMOF and FBLN5

As both FMOD and FBLN5 plasma levels were found to be significantly associated with outcome, we have analyzed what cut-off value divided the patients the best in terms of outcome. ROC AUC analysis showed that the best cut-off value was 27 ng/mL for FMOD and 54 ng/mL for FBLN5 **(Supplementary figure 3**). Patients with elevated FMOD (i.e. > 27 ng/mL, n= 118) or FBLN5 (i.e. > 54 ng/mL, n = 174) had worse event-free survival than the rest of the cohort (**Figure 5 A and B**). Further, we have divided the clinical cohort in three groups according to FMOD and FBLN5 plasma levels as follows: low FMOD/low FBLN5 (n= 40), low FMOD/ high FBLN5 (n= 73) OR high FMOD/low FBLN5 (n=18) and high FMOD/high FBLN5 (n= 100). Patients with the elevated level of one biomarker had significantly worse outcome than those with low level of both biomarkers. Similarly, patients with an elevation of both biomarkers had significantly worse outcome than those with only one elevate biomarker (**Figure 5C**).

**Figure 5:**
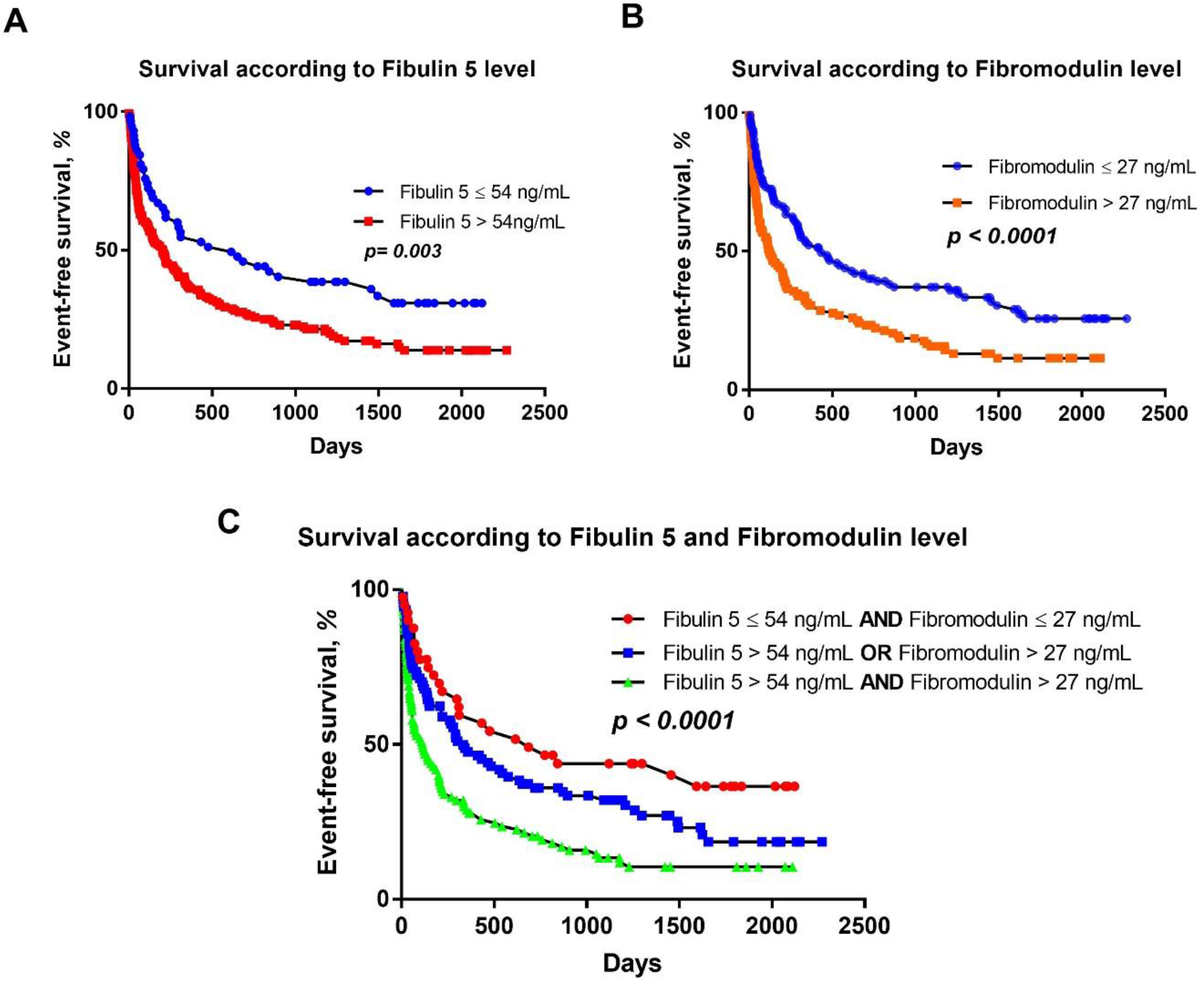
Survival analysis according to the plasma levels of FMOD and FBLN5. Patients were divided according to the plasma concentrations of FMOD and FBLN5. A- Kaplan-Meier analysis showing that patients with FBLN5 concentration > 54 ng/ml had worse outcome than the rest of the cohort, B- Kaplan-Meier analysis showing that patients with FMOD plasma concentration > 27 ng/ml had worse outcome than the rest of the cohort, C- Kaplan-Meier analysis showing the additive prognostic effect when both markers were elevated.

## DISCUSSION

RV dysfunction (RVD) is a commonly accompanying condition in HFrEF patients, and is associated with poor prognosis (3, 11). Unfortunately, no specific biomarker of RVD in HF context is available for clinical routine. In our study, we show that highly sensitive and unbiased proteomic methods can assist identification of the candidate marker proteins, and that circulating concentrations of ECM proteins FMOD and FBL5 may serve as indicators of RV dysfunction and predict outcome of HFrEF patients.

RVD is associated with progressive changes in tissue composition of right ventricle with cardiomyocyte hypertrophy, stress-induced changes in sarcomeric proteins, inflammatory cell infiltration and changes in ECM (3). However, RVD may also specifically affect composition of the left ventricle. Our proteomic approach identified distinct sets of 12 proteins that were differentially expressed in the right ventricles and 6 proteins differentially expressed in the left ventricles of HFrEF patients with severe RVD compared to HFrEF patient without RVD. Among thirteen upregulated proteins six are the component of ECM - FMOD, FBLN5, versican (VCAN), biglycan (BGN), microfibril-associated glycoprotein 4 (MFAP4) and thrombospondin-4 (THBS4), according to their G.O. annotations. Three additional upregulated proteins, namely ubiquitin carboxyl-terminal hydrolase isozyme L1 (UCHL1), non-muscle myosin-10 (MYH10) and C-type mannose receptor 2 (MRC2) are intracellular but contribute to ECM remodeling (6–8).

The results of our proteomic analysis thus clearly suggest that in HFrEF patients, RVD is associated with significant alteration in ECM composition in both ventricles. Similarly to our results, another ECM protein - latent transforming growth factor beta binding protein 2 (LTBP-2) was recently identified using similar approach as a candidate marker of RV dysfunction in patients with pulmonary arterial hypertension (12), confirming relevance of RV extracellular matrix remodelling in the disease progression. Extracellular matrix proteins FMOD and FBLN5 were among the most markedly upregulated proteins in the right and left ventricles, respectively, in HFrEF patients with RVD.

### ECM remodeling and fibrosis in HF

Presence of regenerative, interstitial and perivascular cardiac fibrosis is a key pathologic feature of HF contributing to systolic and diastolic impairment (13, 14) including RVD (12, 15), and has been extensively discussed as a potential therapeutic target (16, 17). The fibrotic ECM remodeling is characterized by extensive production, turnover and crosslinking of collagen fibers orchestrated by numerous matricellular proteins. Five of those have been identified as upregulated in our study (FMOD, BGN, VCAN, MFAP4 and THBS4).

**Fibromodulin (FMOD)** (upregulated in RV of patients with RVD) is a small leucine rich proteoglycan that binds collagen and regulates its crosslinking and collagen fibril organization (18). In vitro, both cardiomyocytes and fibroblasts can produce fibromodulin upon pro-inflammatory stimuli, suggesting that both cell types may participate directly or indirectly in the cardiac ECM remodeling (19). FMOD together with ECM proteoglycans BGN and VCAN (both also upregulated in our study) were previously found to be upregulated in the LV of hearts of patients with ischemic HF, compared to non-failing donor hearts (20). Transcriptomic profiling showed an increased expression of *FMOD* gene in RVD due to pulmonary hypertension (21). Fibromodulin protein upregulation has been recently reported in failing RV due to pulmonary arterial hypertension (12).

Along with collagen fibrils, elastic fibers are an important component of ECM. Formed mainly by proteins elastin and fibrillin, elastic fibers provide tissues, especially lungs, skin and blood vessels, with elasticity in response to mechanical forces (22). In addition, elastic fiber proteins play a role in wound healing, especially in the inflammation phase of the process (22, 23).

**Fibulin-5** (FBLN5) (upregulated in LV in our study) modulates elastic fiber formation. FBLN5 associates with tropoelastin, and assists its deposition into fibrillin-based microfibrils, forming the elastic fibers (22). FBLN5 is essential for elastogenesis and involved in several pathologies including cutis laxa as shown by experimental stuides (24, 25). Expression of FBLN5 is, similarly to collagen fibrillogenesis, regulated by TGFbeta signalling (23, 26). FBLN5 was previously found to be upregulated in LV of patients with ischemic HF compared to non-failing donor hearts (20).

### Fibromodulin and fbulin-5 as biomarkers of RV dysfunction

FMOD and FBLN5 were the most markedly upregulated ECM proteins associated with RVD in HFrEF patients. Their significant upregulation was ventricle-specific, FMOD was upregulated in RV while FBLN5 in LV. As both proteins are secreted/ECM molecules, they were detectable in blood plasma of HFrEF patients as well as healthy controls. However, it should be noted, that other tissues may contribute to the FMOD and FBLN5 plasma pools since the expression of FMOD and FBLN5 is not restricted to the heart.

Nevertheless, plasma concentrations of FMOD and FBLN5 significantly positively correlated with the degree of RV dysfunction, but there was no association of FMOD or FBLN5 plasma levels with LVEF, BMI or kidney function (eGFR). Additionally, plasma levels of both proteins were significantly associated with patient outcome in Cox proportional hazard model suggesting that these proteins mirror pathophysiologically relevant processes with clinical significance.

As the plasma levels of both FMOD and FBLN5 correlated with the degree of RV dysfunction, we have tested whether these proteins had any additional prognostic role beyond RV function alone. Addition of FMOD plasma levels did not improve the RVD-based prognostic information in contrast to FBLN5 which showed an additive prognostic role on top of RV function assessment. This may suggest that increased plasma levels of FBLN5 (upregulated in LV) reflects a pathophysiological process not fully captured by echocardiographic assessment of RVD.

Prognosis of HFrEF patients in clinical settings is driven by the degree of HF itself (LVEF, HF duration, NYHA functional class), the quality of HF treatment (drug/device therapy) as well as associated comorbidities (renal function, presence of diabetes/COPD) (5). MAGGIC score encompassing above-mentioned parameters together with age, blood pressure, BMI, sex and smoking status represents robust and validated tool to estimate the prognosis of HF patients (10). However, MAGGIC composite score does not take into the account RV function. Conversely to the prediction based on RVD, addition of FMOD (upregulated in the RV), but not FBLN5 (upregulated in the LV) plasma levels to MAGGIC score showed an improvement in prognostic assessment compared to MAGGIC score alone. This suggests that increased plasma levels of FMOD reflects a pathophysiological process not addressed by parameters including in the MAGGIC score, potentially a process taking place in the right ventricle, where FMOD was found to be significantly upregulated.

More importantly, the role of FMOD and FBLN5 in prognostic HFrEF assessment was additive. As HF is a complex syndrome affecting multiple tissues, vascular ECM remodeling or pro-fibrotic changes in lungs, kidney, liver or spleen of HF patients may also theoretically increase the FBLN5 a FMOD plasma concentrations, and contribute to the prognostic value of the proteins in HFrEF patients. Nevertheless, the additivity may still be attributed to the distinct pathophysiological roles of the proteins (i.e. collagen deposition/ crosslinking for FMOD, elastic fiber formation for FBLN5) or to a different activity of the processes in the respective ventricles. These properties make FMOD and FBLN5 promising blood biomarkers integrating multiple HF-related processes in addition to RV dysfunction.

## Conclusions

Increased myocardial expression of extracellular matrix proteins FMOD and FBLN5 was identified in HFrEF patients with RVD. Both proteins were detectable in plasma in both HFrEF patients as well as HF-free individuals, their plasma levels positively correlated with the degree of RVD, and both were associated with an adverse outcome. They provided additional prognostic information beyond already established parameters (RV function assessment, MAGGIC score) and their prognostic information was additive. Our study thus proposes plasma levels of FMOD and FBLN5 as new biomarkers for HFrEF patients reaching beyond RV dysfunction only.

## Limitations

We cannot conclude whether the observed FMOD and FBLN5 upregulation in myocardium of HFrEF patients with RVD is to be attributed to cardiomyocytes, fibroblasts or other cell types present in the myocardium. Similarly, we cannot determine whether myocardium is the only tissue responsible for the increased circulating concentrations of the proposed biomarkers.

Patients in our study were treated not only by optimal pharmacotherapy/ICD/CRT device implantation, but some of them underwent heart transplantation or implantation of mechanical circulatory support, which may bias the outcome analysis. As the patients were enrolled before SGL2i era, none was treated with these drugs. Although patients were prospectively enrolled, not all of them were followed in our hospital; therefore the information about changes in pharmacotherapy throughout the follow-up period is not available from all the patients. Similarly, data about further cardiac decompensation events were not available from all the patients, so it was not possible to analyze this endpoint. Our study cohort included rather young patients with more advanced HF; consequently, the results might not be fully applicable to patients with milder HF or to older patients. Similarly, whether FMOL or FBLN5 reflect the degree of RV dysfunction in HFmrEF or HFpEF patients cannot be concluded from our study. As we have strived for maximal applicability of our results, we have included all HF patients regardless of etiology. However, differences in ECM remodeling may exist depending upon HF etiology.

## Disclosure

Authors declare no conflict of interest

## Data Availability

All data are availabe on reasonable request. The mass spectrometry data have been deposited to the ProteomeXchange Consortium via the PRIDE partner repository with the dataset identifier PXD043768. Before publication, reviewers may access the data via following login and password: Username: Password: XWgGXHiw

https://proteomecentral.proteomexchange.org/cgi/GetDataset?ID=PXD043768

